# Poor Sleep Quality is associated with Decreased Brain Glucose Metabolism in Healthy Middle-aged Adults

**DOI:** 10.1101/2024.04.29.24306506

**Authors:** Seunghyeon Shin, Ju Won Seok, Keunyoung Kim, Jihyun Kim, Hyun-Yeol Nam, Kyoungjune Pak

## Abstract

Sleep disturbance is associated with the development of neurodegenerative disease. We aimed to address the effects of sleep quality on brain glucose metabolism measured by ^18^F-Fluorodeoxyglucose (^18^F-FDG) positron emission tomography (PET) in healthy middle-aged adults. A total of 378 healthy men (mean age: 42.8±3.6 years) were included in this study. Participants underwent brain ^18^F-FDG PET and completed the Korean version of the Pittsburgh Sleep Quality Index (PSQI-K). Additionally, anthropometric measurements were obtained. PETs were spatially normalized to MNI space using PET templates from SPM5 with PMOD. The Automated Anatomical Labeling 2 atlas was used to define regions of interest (ROIs). The mean uptake of each ROI was scaled to the mean of the global cortical uptake of each individual and defined as the standardized uptake value ratio (SUVR). After the logarithmic transformation of the regional SUVR, the effects of the PSQI-K on the regional SUVR were investigated using Bayesian hierarchical modeling. Brain glucose metabolism of the posterior cingulate, precuneus, and thalamus showed a negative association with total PSQI-K scores in the Bayesian model ROI-based analysis. Voxel-based analysis using statistical parametric mapping revealed a negative association between the total PSQI-K scores and brain glucose metabolism of the precuneus, postcentral gyrus, posterior cingulate, and thalamus. Poor sleep quality is negatively associated with brain glucose metabolism in the precuneus, posterior cingulate, and thalamus. This finding may provide a link between sleep quality and the risk of neurodegenerative disease. Therefore, the importance of sleep should not be overlooked, even in healthy middle-aged adults.

## 1. INTRODUCTION

The average person spends more than 25 years sleeping in their lifetime. Insufficient sleep is associated with obesity, diabetes, depression, memory, and learning difficulty (Albakri et al., 2021), and higher sleep quality is associated with increased cognitive test performance (Gildner et al., 2014). Moreover, sleep disturbance increases the risk of dementia (Sabia et al., 2021; Shi et al., 2018) and is frequent in neurodegenerative diseases, including Alzheimer’s disease (AD) (Pak et al., 2020). Thus, sleep disturbance is not only a symptom of underlying neurodegenerative disease but is also a contributing factor in the development and progression of neurodegenerative disease (Abbott and Videnovic, 2016). Sleeplessness causes metabolic byproducts to accumulate, and the amyloid beta (Aβ), the pathologic hallmark of AD (Brown et al., 2014), is cleared by the glymphatic pathway during sleep (Pak et al., 2020). Even one night of sleep deprivation can lead to increased amyloid deposition in the human brain, as confirmed by amyloid measurements using positron emission tomography/computed tomography (PET/CT) (Shokri-Kojori et al., 2018).

The human brain utilizes glucose as its primary source of energy; thus, brain glucose metabolism, assessed by PET with ^18^F-Fluorodeoxyglucose (FDG), can be utilized for quantifying neuronal activity in the human brain (de Leon et al., 2001). ^18^F-FDG PET/CT is a useful imaging modality for the diagnosis, differentiation of dementia, and prediction of mild cognitive impairment leading to AD (Brown et al., 2014). Several studies with inconsistent results investigated the association between sleep quality and brain glucose metabolism in late adulthood (Branger et al., 2016; Kimura et al., 2020; Stankeviciute et al., 2023). Stankeviciute et al. reported negative associations between the Pittsburgh Sleep Quality Index (PSQI), a measure of sleep quality, and brain glucose metabolism in the right temporal pole, right paracingulate gyrus, right cerebellum exterior, and right frontal orbital cortex (Stankeviciute et al., 2023). No association between PSQI and brain glucose metabolism was observed in a study by Branger et al. (Branger et al., 2016). Kimura et al. demonstrated that total sleep time was inversely associated with brain glucose metabolism but not sleep efficiency (Kimura et al., 2020). These studies enrolled older adults with or without cognitive impairment. Aging was found to be associated with decreased brain glucose metabolism and increased amyloid burden (Pourhassan Shamchi et al., 2018; Rodrigue et al., 2012). A longitudinal study reported that decreased brain glucose metabolism was associated with aging by identifying this trend in cognitively normal older adults (Ishibashi et al., 2018). Moreover, several studies reported that brain glucose metabolism interacted with Aβ and tau protein in older adults (Crook et al., 2021; Hanseeuw et al., 2017). Thus, aging, Aβ, and tau protein could interfere with the effect of sleep disturbance on brain glucose metabolism in older adults. If the effect of sleep quality on the brain is present, this would be identified even in middle-aged adults without the effect of aging, Aβ, and tau protein.

As the risk of AD increases, especially after the age of 65 years, the importance of sleep is overlooked in middle adulthood. In addition, aging itself leads to the decline of brain glucose metabolism in the caudate, cingulate, frontal, and parietal lobes, according to our previous study with ^18^F-FDG PET (Pak et al., 2023). Therefore, to address the effects of sleep quality on brain glucose metabolism, we analyzed a large cohort of healthy middle-aged adults who underwent brain ^18^F-FDG PET and completed a sleep quality questionnaire. We used Bayesian hierarchical modeling to estimate the effects of sleep quality on brain glucose metabolism and hypothesized that poor sleep quality is negatively associated with brain glucose metabolism.

## 2. MATERIALS AND METHODS

### 2.1. Participants

We retrospectively analyzed data from 473 healthy men who participated in the health checkup program at the Samsung Changwon Hospital Health Promotion Center in 2013. After excluding individuals with neuropsychiatric disorders (n=5) or malignancies (n=3), those with missing data from the Korean version of PSQI (PSQI-K) (n=87) (Sohn et al., 2012), 378 healthy men were included. The health checkup program included 1) brain ^18^F-FDG PET, 2) anthropometric measurements, and 3) completion of the PSQI-K. Participants in this study were included in a previous study on the effect of aging on brain glucose metabolism (Pak et al., 2023). The study protocol was approved by the Institutional Review Board of Changwon Samsung Hospital. The requirement for informed consent was waived owing to the retrospective study design.

### 2.2. Brain ^18^F-FDG PET and image analysis

The participants were asked to avoid strenuous exercise for 24 hours and to fast for at least 6 hours before the PET study. PET/CT was performed 60 mins after injection of ^18^F-FDG (3.7 MBq/kg) with the Discovery 710 PET/CT scanner (GE Healthcare, Waukesha, WI, USA). Continuous spiral CT was obtained with a tube voltage of 120 kVp and tube current of 30–180 mAs. PET was obtained in three-dimensional mode with full width at half maximum of 5.6 mm and reconstructed using an ordered-subset expectation maximization algorithm. PETs were spatially normalized to MNI space using PET templates from SPM5 (University College of London, UK) with PMOD version 3.6 (PMOD Technologies LLC, Zurich, Switzerland). Automated Anatomical Labeling 2 atlas (Rolls et al., 2015) was used to define regions of interest (ROIs): caudate, putamen, thalamus, cingulate (anterior, middle, posterior), frontal (middle, superior), hippocampus, parietal (inferior, superior), postcentral, precuneus, temporal (inferior, middle, superior) gyri, and cerebellum. The mean uptake of each ROI was scaled to the mean of the global cortical uptake of each individual and defined as the standardized uptake value ratio (SUVR). For a full-volume analysis, the statistical threshold was set at a cluster level and corrected with a false discovery rate with p < 0.05 in a regression model (correction with age) after smoothing SUVR images with a Gaussian kernel of full width at half maximum 8mm (Statistical Parametric Mapping 12, Wellcome Centre for Human Neuroimaging, UCL, London, UK).

### 2.3. PSQI-K

All participants completed the PSQI-K to assess subjective quality and sleep patterns (Sohn et al., 2012). The PSQI-K consists of 19 self-reported items with seven subcategories: 1) subjective sleep quality, 2) sleep latency, 3) sleep duration, 4) habitual sleep efficiency, 5) sleep disturbances, 6) use of sleep medication, and 7) daytime dysfunction. Scores for each subcategory range between 0 and 3, with higher scores indicating worse sleep quality; total scores can range from 0 to 21.

### 2.4. Statistical analysis

Normality was tested with the Shapiro–Wilk test. After the logarithmic transformation of the regional SUVR, the effects of the PSQI-K on the regional SUVR were investigated using Bayesian hierarchical modeling with brms (Bürkner, 2017, 2018, 2021) that applies the Markov–Chain Monte Carlo sampling tools of RStan (Stan Development Team, 2022). We set up models separately for total PSQI-K scores and subcategories with the regional SUVR as a dependent variable and PSQI-K scores as predictors adjusting for age. These fixed effects (PSQI-K and age) were calculated individually, and participants and ROI were added as random intercepts to allow the SUVR to vary between participants and ROIs. Bayesian models were estimated using four Markov chains, each with 4,000 iterations, including 1,000 warm-ups, thus totaling 12,000 post-warm-up samples. The sampling parameters were slightly modified to facilitate convergence (max treedepth = 20). Statistical analysis was performed in R Statistical Software (The R Foundation for Statistical Computing).

## 3. RESULTS

### 3.1. Participant characteristics

A total of 378 healthy men (mean age: 42.8±3.6 years) were included in this study. The mean and standard deviation of the total score of the PSQI-K was 4.0±2.2, ranging from 0 to 12. The mean and standard deviation of the scores of the seven PSQI-K subcategories are as follows: 1) subjective sleep quality; 1.0±0.6, 2) sleep latency; 0.8±0.8, 3) sleep duration; 0.9±0.7, 4) habitual sleep efficiency; 0.2±0.4, 5) sleep disturbances; 0.6±0.5, 6) use of sleep medication; 0±0.1, 7) daytime dysfunction; 0.6±0.6 (Table 1).

**Table 1.** Participant characteristics. PSQI-K, Pittsburgh Sleep Quality Index-Korean version.

|  | Mean±standard deviation |
| --- | --- |
| Age (years) | 42.8±3.6 |
| Body mass index (kg/m<sup>2</sup>) | 24.7±2.9 |
| Total PSQI-K | 4.0±2.2 |
| 1) subjective sleep quality | 1.0±0.6 |
| 2) sleep latency | 0.8±0.8 |
| 3) sleep duration | 0.9±0.7 |
| 4) habitual sleep efficiency | 0.2±0.4 |
| 5) sleep disturbances | 0.6±0.5 |
| 6) use of sleep medication | 0±0.1 |
| 7) daytime dysfunction | 0.6±0.6 |

### 3.2. Sleep quality and brain glucose metabolism

Brain glucose metabolism of the posterior cingulate, precuneus, and thalamus showed a negative association with total PSQI-K scores (Figure 1). Full-volume analysis revealed a consistent negative association of total PSQI-K scores with brain glucose metabolism of the precuneus, postcentral gyrus, posterior cingulate, and thalamus (Figure 2). No area positively correlated with total PSQI-K scores from ROI- and voxel-based analyses.

**Figure 1.**
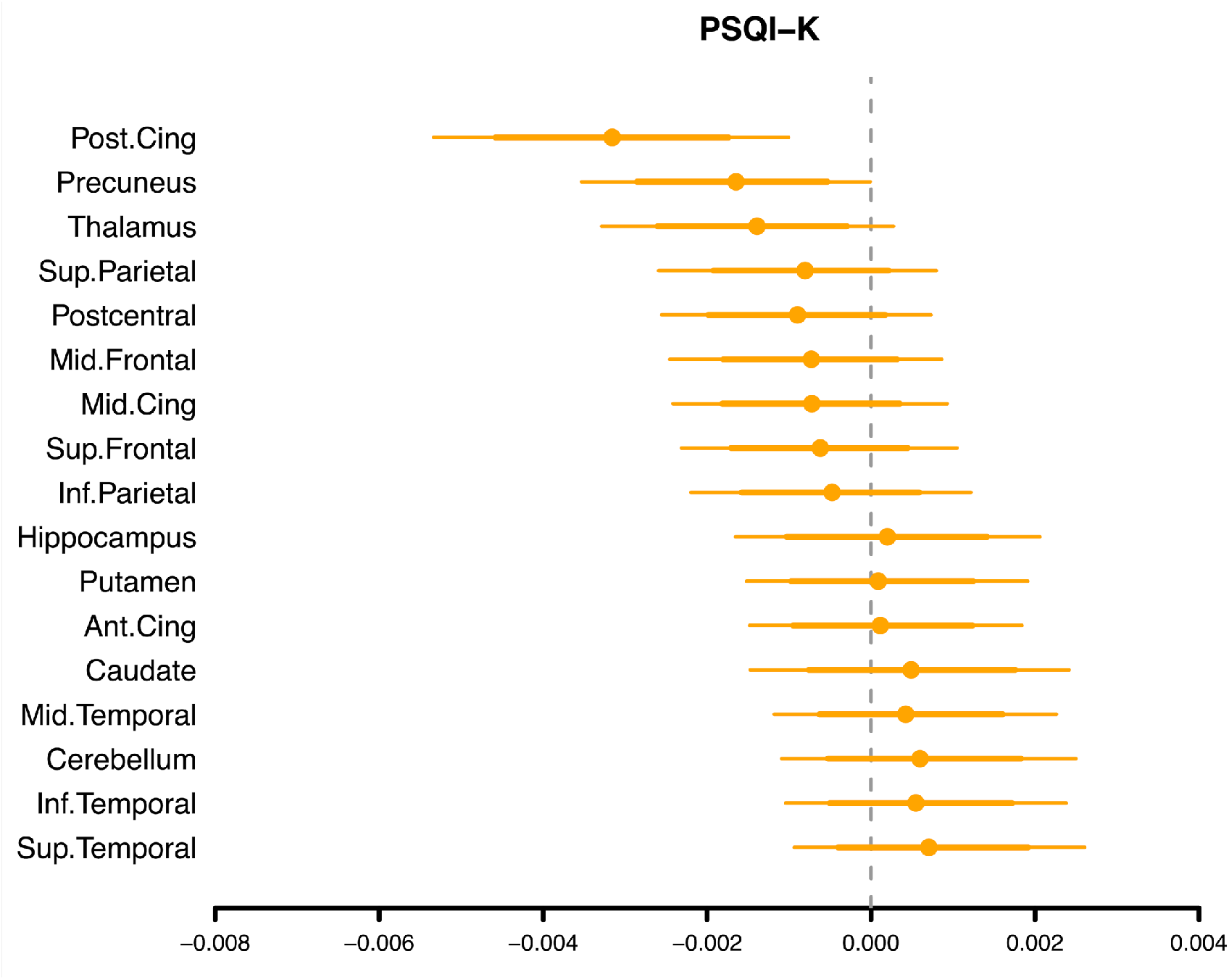
Posterior intervals of the regression coefficients for total PSQI-K scores predicting brain glucose metabolism. The thick lines represent the 80% posterior intervals, the thin lines represent the 95% posterior intervals, and the circles represent the posterior means. PSQI-K, Pittsburgh Sleep Quality Index-Korean version.

**Figure 2.**
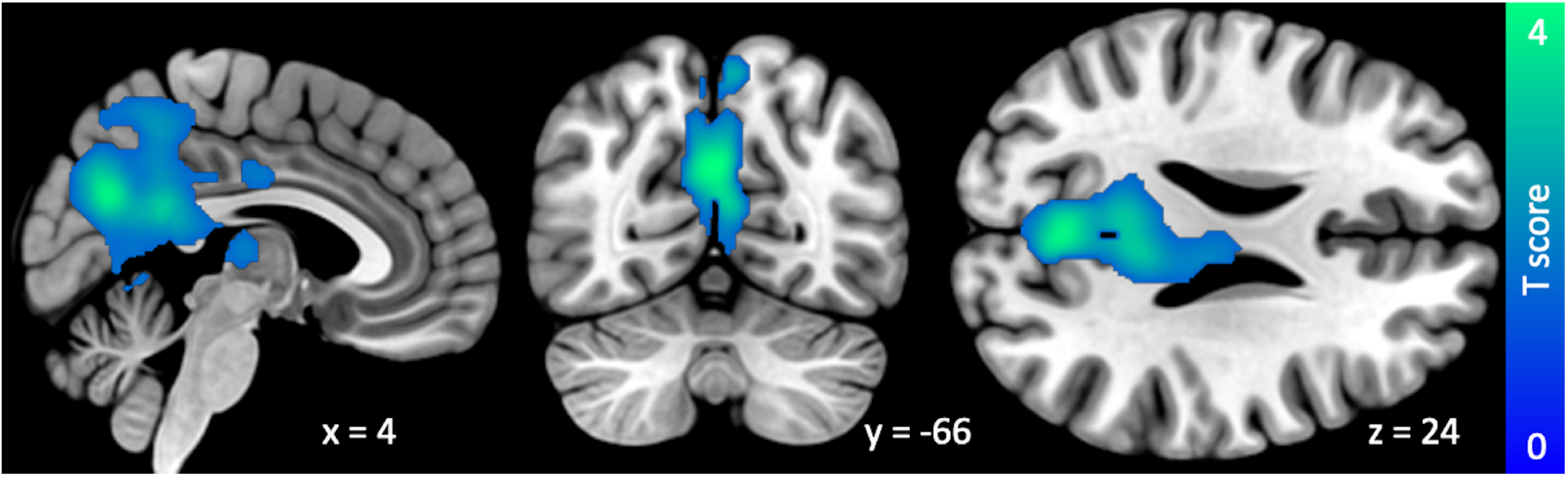
Full volume analysis showing the negative association of total PSQI-K scores with brain glucose metabolism. PSQI-K, Pittsburgh Sleep Quality Index-Korean version.

In Bayesian models with PSQI-K subcategories, brain glucose metabolism of the posterior cingulate and precuneus showed a negative association with the scores of PSQI subcategories (subjective sleep quality, sleep latency, sleep duration, use of sleep medication, and daytime dysfunction) with some of their 95% posterior intervals overlapping with zero. In models using sleep medication and daytime dysfunction, poor sleep quality was associated with decreased brain glucose metabolism across the regions, with some of their 95% posterior intervals overlapping with zero (Figure 3).

**Figure 3.**
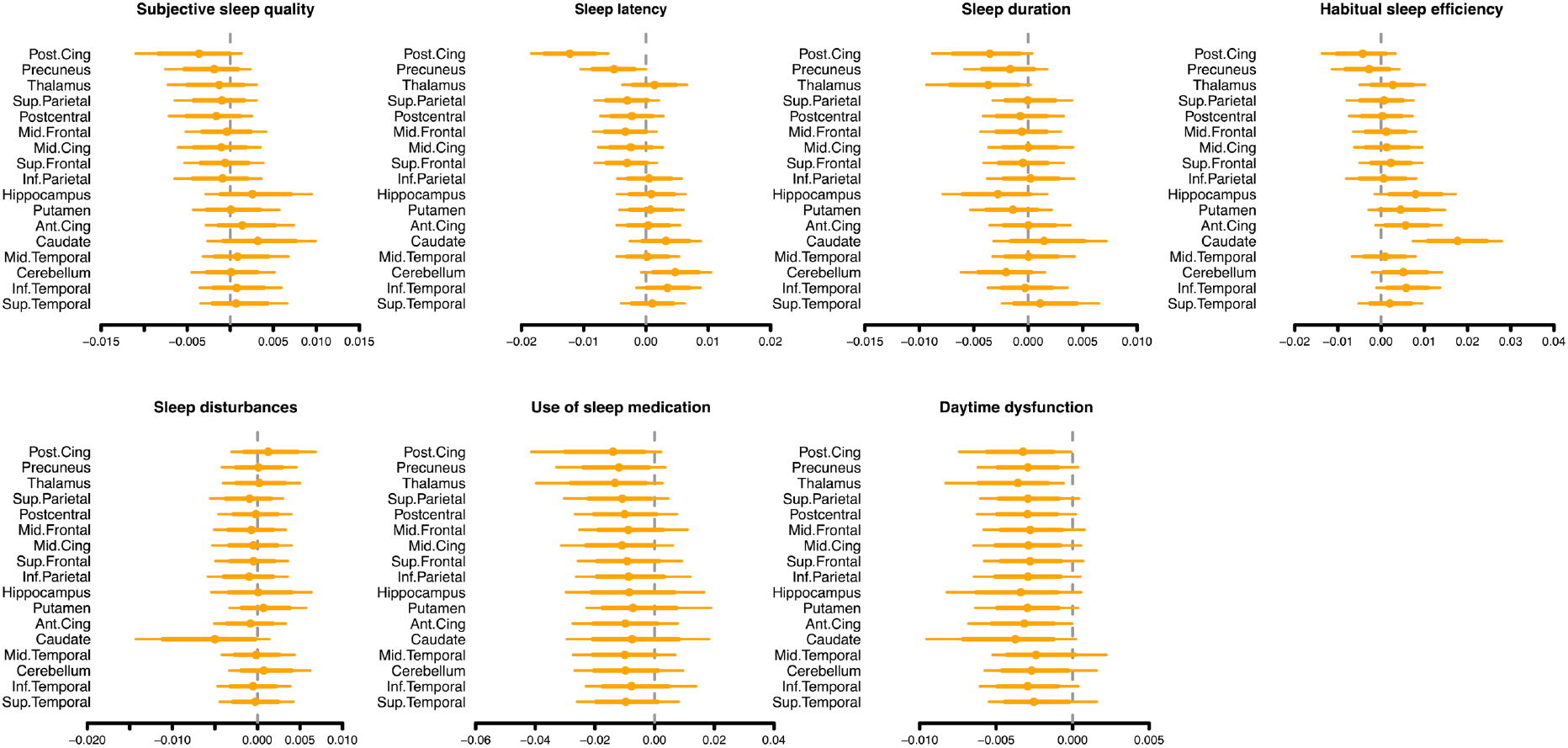
Posterior intervals of the regression coefficients for the subcategories of the PSQI-K predicting brain glucose metabolism. The thick lines represent the 80% posterior intervals, the thin lines represent the 95% posterior intervals, and the circles represent the posterior means. PSQI-K, Pittsburgh Sleep Quality Index-Korean version.

## 4. DISCUSSION

Our main finding was that poor sleep quality measured by the PSQI-K of 378 middle-aged men (mean age: 42.8±3.6 years) was negatively associated with brain glucose metabolism in the precuneus, posterior cingulate, and thalamus in ROI- and voxel-based analyses.

AD is the most common cause of dementia and a slowly progressive neurodegenerative disease (Abubakar et al., 2022). Cognitive decline in AD is associated with the accumulation of the Aβ (Abubakar et al., 2022), and this is associated with an imbalance between Aβ neuronal production and extracellular clearance of Aβ (Hampel et al., 2021). Aβ is identified by a cerebrospinal fluid Aβ42 assay and amyloid PET before the onset of clinical symptoms (Hampel et al., 2021). Also, brain ^18^F-FDG PET can demonstrate a decrease in brain glucose metabolism, indicative of synaptic dysfunction, in the parietotemporal cortex, posterior cingulate, and precuneus in the early stages of AD (Hampel et al., 2021).

Sleep disturbance increases the risk of dementia (Sabia et al., 2021; Shi et al., 2018) and is frequent in neurodegenerative diseases, including AD (Pak et al., 2020). One night of sleep deprivation can cause increased amyloid burden in healthy adults, according to amyloid PET studies (Shokri-Kojori et al., 2018). Sleep disturbance and short sleep duration are also associated with poorer cognitive function (Behrens et al., 2023; Lo et al., 2016). The classic AD pattern in brain ^18^F-FDG PET is hypometabolism in the parietotemporal cortex, posterior cingulate, and precuneus (Brown et al., 2014), which are associated with cognitive decline, including memory and attention (Cavanna and Trimble, 2006; Leech and Sharp, 2014). Thus, the association between the PSQI-K and brain glucose metabolism may indicate a link between poor sleep quality and AD, even in middle adulthood.

In addition, a structural magnetic resonance imaging (MRI) study revealed that poor sleep quality was associated with reduced volume of the right superior frontal cortex and increased rate of atrophy of the frontal, temporal, and parietal lobes in individuals over 60 years (Sexton et al., 2014). Other MRI studies also demonstrated the association between sleep disturbance and decreased gray matter volume (Alperin et al., 2019; Branger et al., 2016; Chao et al., 2014; Li et al., 2019; Park et al., 2020; Stankeviciute et al., 2023). Therefore, these results may also reflect the association between sleep quality and structural changes in brain regions. Sleeplessness can cause metabolic byproducts to accumulate, and the Aβ, the pathologic hallmark of AD (Brown et al., 2014), is cleared by the glymphatic pathway during sleep (Pak et al., 2020). Even one night of sleep deprivation leads to increased amyloid deposition in the hippocampal, parahippocampal, and thalamic regions of the human brain, according to amyloid PET studies (Shokri-Kojori et al., 2018; Spira et al., 2013). Also, shorter sleep duration was associated with higher amyloid burden in the bilateral putamen, parahippocampus, and right precuneus after sleep (Shokri-Kojori et al., 2018).

Patients with primary insomnia had lower brain glucose metabolism in the precuneus, posterior cingulate cortex, anterior cingulate, medial frontal cortex, right hippocampus/amygdala, and right fusiform gyrus (Kay et al., 2016). Patients with sleep apnea, associated with sleep fragmentation and risk of cognitive impairment, had lower brain glucose metabolism in the precuneus, posterior cingulate cortex, and frontal area, probably due to nocturnal hypoxia and sleep fragmentation (Fernandes et al., 2022).

Recently, a negative association between the PSQI and brain glucose metabolism in the right temporal pole, right paracingulate gyrus, right cerebellum exterior, and right frontal orbital cortex was reported in a study with participants with an average age of 61.2 years (Stankeviciute et al., 2023). However, another study showed no association between sleep quality and brain glucose metabolism in participants with an average of 64.1 years of age (Branger et al., 2016).

This study included 378 healthy men with a mean age of 42.8 years. Even in middle adulthood, sleep quality was negatively associated with brain glucose metabolism of the precuneus, posterior cingulate, and thalamus. In an MRI study including adults with a mean age of 46.4 years, no association was observed between sleep quality and grey matter volume (Hidese et al., 2023). Sleep quality was associated with a decline in grey matter volume of the frontal, temporal, and parietal lobes in participants over 60 but not in those under 60 years (Sexton et al., 2014). Therefore, the decline of brain glucose metabolism associated with sleep quality may be dependent on age and further accelerated in the older population.

This study has several limitations. Only men were included in this study; therefore, the results may not be generalizable and directly applicable to women. This retrospective study was based on a health checkup program. As a brain MRI was not included in the program, MRI-based coregistration and partial volume correction of PETs could not be done, and the results could not be compared with MR-based indices of atrophy.

## 5. CONCLUSION

Sleep quality is negatively associated with brain glucose metabolism in the precuneus, posterior cingulate, and thalamus. This finding may indicate an association between sleep quality and the risk of neurodegenerative diseases, such as AD. Therefore, the importance of sleep should not be overlooked in middle-aged healthy adults.

## Data Availability

All data produced in the present study are available upon reasonable request to the authors

## ACKNOWLEDGEMENT

None

## REFERENCES

Abbott, S.M., Videnovic, A., 2016. Chronic sleep disturbance and neural injury: links to neurodegenerative disease. Nat Sci Sleep 8, 55–61.

Abubakar, M.B., Sanusi, K.O., Ugusman, A., Mohamed, W., Kamal, H., Ibrahim, N.H., Khoo, C.S., Kumar, J., 2022. Alzheimer’s Disease: An Update and Insights Into Pathophysiology. Front Aging Neurosci 14, 742408.

Albakri, U., Drotos, E., Meertens, R., 2021. Sleep Health Promotion Interventions and Their Effectiveness: An Umbrella Review. Int J Environ Res Public Health 18.

Alperin, N., Wiltshire, J., Lee, S.H., Ramos, A.R., Hernandez-Cardenache, R., Rundek, T., Curiel Cid, R., Loewenstein, D., 2019. Effect of sleep quality on amnestic mild cognitive impairment vulnerable brain regions in cognitively normal elderly individuals. Sleep 42.

Behrens, A., Anderberg, P., Berglund, J.S., 2023. Sleep disturbance predicts worse cognitive performance in subsequent years: A longitudinal population-based cohort study. Arch Gerontol Geriatr 106, 104899.

Branger, P., Arenaza-Urquijo, E.M., Tomadesso, C., Mezenge, F., Andre, C., de Flores, R., Mutlu, J., de La Sayette, V., Eustache, F., Chetelat, G., Rauchs, G., 2016. Relationships between sleep quality and brain volume, metabolism, and amyloid deposition in late adulthood. Neurobiol Aging 41, 107–114.

Brown, R.K., Bohnen, N.I., Wong, K.K., Minoshima, S., Frey, K.A., 2014. Brain PET in suspected dementia: patterns of altered FDG metabolism. Radiographics 34, 684–701.

Bürkner, P.-C., 2017. brms: an R package for Bayesian multilevel models using Stan. Journal of Statistical Software 80, 1–28.

Bürkner, P.-C., 2018. Advanced Bayesian multilevel modeling with the R package brms. The R Journal 10, 395–411.

Bürkner, P.-C., 2021. Bayesian Item Response Modeling in R with brms and Stan. Journal of Statistical Software 100, 1–54.

Cavanna, A.E., Trimble, M.R., 2006. The precuneus: a review of its functional anatomy and behavioural correlates. Brain 129, 564–583.

Chao, L.L., Mohlenhoff, B.S., Weiner, M.W., Neylan, T.C., 2014. Associations between subjective sleep quality and brain volume in Gulf War veterans. Sleep 37, 445–452.

Crook, H., Nowell, J., Raza, S., Edison, P., 2021. Longitudinal FDG-PET and FTP-PET demonstrates the relationship between glucose metabolism and tau accumulation in Alzheimer’s disease atrophy-associated brain regions in cognitively normal older adults. Alzheimer’s & Dementia 17, e056276.

de Leon, M.J., Convit, A., Wolf, O.T., Tarshish, C.Y., DeSanti, S., Rusinek, H., Tsui, W., Kandil, E., Scherer, A.J., Roche, A., Imossi, A., Thorn, E., Bobinski, M., Caraos, C., Lesbre, P., Schlyer, D., Poirier, J., Reisberg, B., Fowler, J., 2001. Prediction of cognitive decline in normal elderly subjects with 2-[(18)F]fluoro-2-deoxy-D-glucose/poitron-emission tomography (FDG/PET). Proc Natl Acad Sci U S A 98, 10966–10971.

Fernandes, M., Mari, L., Chiaravalloti, A., Paoli, B., Nuccetelli, M., Izzi, F., Giambrone, M.P., Camedda, R., Bernardini, S., Schillaci, O., Mercuri, N.B., Placidi, F., Liguori, C., 2022. (18)F-FDG PET, cognitive functioning, and CSF biomarkers in patients with obstructive sleep apnoea before and after continuous positive airway pressure treatment. J Neurol 269, 5356–5367.

Gildner, T.E., Liebert, M.A., Kowal, P., Chatterji, S., Snodgrass, J.J., 2014. Associations between sleep duration, sleep quality, and cognitive test performance among older adults from six middle income countries: results from the Study on Global Ageing and Adult Health (SAGE). J Clin Sleep Med 10, 613–621.

Hampel, H., Hardy, J., Blennow, K., Chen, C., Perry, G., Kim, S.H., Villemagne, V.L., Aisen, P., Vendruscolo, M., Iwatsubo, T., Masters, C.L., Cho, M., Lannfelt, L., Cummings, J.L., Vergallo, A., 2021. The Amyloid-beta Pathway in Alzheimer’s Disease. Mol Psychiatry 26, 5481–5503.

Hanseeuw, B.J., Betensky, R.A., Schultz, A.P., Papp, K.V., Mormino, E.C., Sepulcre, J., Bark, J.S., Cosio, D.M., LaPoint, M., Chhatwal, J.P., Rentz, D.M., Sperling, R.A., Johnson, K.A., 2017. Fluorodeoxyglucose metabolism associated with tau-amyloid interaction predicts memory decline. Ann Neurol 81, 583–596.

Hidese, S., Ota, M., Matsuo, J., Ishida, I., Yokota, Y., Hattori, K., Yomogida, Y., Kunugi, H., 2023. Association between the Pittsburgh sleep quality index and white matter integrity in healthy adults: a whole-brain magnetic resonance imaging study. Sleep and Biological Rhythms 21, 249–256.

Ishibashi, K., Onishi, A., Fujiwara, Y., Oda, K., Ishiwata, K., Ishii, K., 2018. Longitudinal effects of aging on (18)F-FDG distribution in cognitively normal elderly individuals. Sci Rep 8, 11557.

Kay, D.B., Karim, H.T., Soehner, A.M., Hasler, B.P., Wilckens, K.A., James, J.A., Aizenstein, H.J., Price, J.C., Rosario, B.L., Kupfer, D.J., Germain, A., Hall, M.H., Franzen, P.L., Nofzinger, E.A., Buysse, D.J., 2016. Sleep-Wake Differences in Relative Regional Cerebral Metabolic Rate for Glucose among Patients with Insomnia Compared with Good Sleepers. Sleep 39, 1779–1794.

Kimura, N., Aso, Y., Yabuuchi, K., Ishibashi, M., Hori, D., Sasaki, Y., Nakamichi, A., Uesugi, S., Jikumaru, M., Sumi, K., Eguchi, A., Obara, H., Kakuma, T., Matsubara, E., 2020. Association of Modifiable Lifestyle Factors With Cortical Amyloid Burden and Cerebral Glucose Metabolism in Older Adults With Mild Cognitive Impairment. JAMA Netw Open 3, e205719.

Leech, R., Sharp, D.J., 2014. The role of the posterior cingulate cortex in cognition and disease. Brain 137, 12–32.

Li, M., Wang, R., Zhao, M., Zhai, J., Liu, B., Yu, D., Yuan, K., 2019. Abnormalities of thalamus volume and resting state functional connectivity in primary insomnia patients. Brain Imaging Behav 13, 1193–1201.

Lo, J.C., Groeger, J.A., Cheng, G.H., Dijk, D.J., Chee, M.W., 2016. Self-reported sleep duration and cognitive performance in older adults: a systematic review and meta-analysis. Sleep Med 17, 87–98.

Pak, K., Kim, J., Kim, K., Kim, S.J., Kim, I.J., 2020. Sleep and Neuroimaging. Nucl Med Mol Imaging 54, 98–104.

Pak, K., Malen, T., Santavirta, S., Shin, S., Nam, H.Y., De Maeyer, S., Nummenmaa, L., 2023. Brain Glucose Metabolism and Aging: A 5-Year Longitudinal Study in a Large Positron Emission Tomography Cohort. Diabetes Care 46, e64–e66.

Park, C.-h., Bang, M., Ahn, K.J., Kim, W.J., Shin, N.-Y., 2020. Sleep disturbance-related depressive symptom and brain volume reduction in shift-working nurses. Scientific Reports 10, 9100.

Pourhassan Shamchi, S., Khosravi, M., Taghvaei, R., Zirakchian Zadeh, M., Paydary, K., Emamzadehfard, S., Werner, T.J., Hoilund-Carlsen, P.F., Alavi, A., 2018. Normal patterns of regional brain (18)F-FDG uptake in normal aging. Hell J Nucl Med 21, 175–180.

Rodrigue, K.M., Kennedy, K.M., Devous, M.D., Jr., Rieck, J.R., Hebrank, A.C., Diaz-Arrastia, R., Mathews, D., Park, D.C., 2012. beta-Amyloid burden in healthy aging: regional distribution and cognitive consequences. Neurology 78, 387–395.

Rolls, E.T., Joliot, M., Tzourio-Mazoyer, N., 2015. Implementation of a new parcellation of the orbitofrontal cortex in the automated anatomical labeling atlas. Neuroimage 122, 1–5.

Sabia, S., Fayosse, A., Dumurgier, J., van Hees, V.T., Paquet, C., Sommerlad, A., Kivimaki, M., Dugravot, A., Singh-Manoux, A., 2021. Association of sleep duration in middle and old age with incidence of dementia. Nat Commun 12, 2289.

Sexton, C.E., Storsve, A.B., Walhovd, K.B., Johansen-Berg, H., Fjell, A.M., 2014. Poor sleep quality is associated with increased cortical atrophy in community-dwelling adults. Neurology 83, 967–973.

Shi, L., Chen, S.J., Ma, M.Y., Bao, Y.P., Han, Y., Wang, Y.M., Shi, J., Vitiello, M.V., Lu, L., 2018. Sleep disturbances increase the risk of dementia: A systematic review and meta-analysis. Sleep Med Rev 40, 4–16.

Shokri-Kojori, E., Wang, G.J., Wiers, C.E., Demiral, S.B., Guo, M., Kim, S.W., Lindgren, E., Ramirez, V., Zehra, A., Freeman, C., Miller, G., Manza, P., Srivastava, T., De Santi, S., Tomasi, D., Benveniste, H., Volkow, N.D., 2018. beta-Amyloid accumulation in the human brain after one night of sleep deprivation. Proc Natl Acad Sci U S A 115, 4483–4488.

Sohn, S.I., Kim, D.H., Lee, M.Y., Cho, Y.W., 2012. The reliability and validity of the Korean version of the Pittsburgh Sleep Quality Index. Sleep Breath 16, 803–812.

Spira, A.P., Gamaldo, A.A., An, Y., Wu, M.N., Simonsick, E.M., Bilgel, M., Zhou, Y., Wong, D.F., Ferrucci, L., Resnick, S.M., 2013. Self-reported sleep and beta-amyloid deposition in community-dwelling older adults. JAMA Neurol 70, 1537–1543.

Stan Development Team, 2022. RStan: the R interface to Stan.

Stankeviciute, L., Falcon, C., Operto, G., Garcia, M., Shekari, M., Iranzo, A., Ninerola-Baizan, A., Perissinotti, A., Minguillon, C., Fauria, K., Molinuevo, J.L., Zetterberg, H., Blennow, K., Suarez-Calvet, M., Cacciaglia, R., Gispert, J.D., Grau-Rivera, O., and for the, A.s., 2023. Differential effects of sleep on brain structure and metabolism at the preclinical stages of AD. Alzheimers Dement.

